# Impact of minimum wage increases on homicide mortality in the US

**DOI:** 10.64898/2026.05.21.26353800

**Authors:** Kate Vinita Fitch, N. Jeanie Santaularia Gomez, Maryam Tanveer, Mark Holmes, Kathryn E. Moracco, Michael D. Fliss, Naoko Fulcher, Shabbar I. Ranapurwala

## Abstract

**Introduction:** Even though state minimum wage (MW) is a policy lever that affects income and poverty and can prevent of violence, no prior study has comprehensively evaluated its impact in the United States (US). In this study, we estimated the impact of at least a $1 USD increase in state MW above the federal MW on overall, firearm, and non-firearm homicide mortality and examined its impact on racialized inequities.

**Methods:** We conducted a quasi-experimental study using controlled interrupted time series (CITS) and synthetic controlled interrupted time series (SCITS) approaches to examine immediate and sustained impact of state MW increases. We used state-month level homicide victimization mortality data from 2010-2019. Homicide victimization death was identified using International Classification of Disease codes, 10^th^ revision. State MW data was obtained from the Bureau of Labor Statistics.

**Results:** Demographic and social variables from intervention, never-exposed, and always-exposed states were similar to each other and representative of the total US population from all 50 states. The CITS results show that after MW increases in the intervention states, these states experienced a sustained decline of −0.22 (−0.37, −0.07) homicide victimizations/ 100,000 person-years/ year relative to the never-exposed states and −0.39 (−0.59, −0.18) relative to always-exposed states. This resulted in 5,657 fewer homicide victimization deaths in the intervention states over four years of post-MW increase period compared to the never-exposed states. SCITS results were similar to the CITS results, and the majority of sustained declines were observed in firearm-related deaths and among Black population.

**Conclusion:** MW increase was associated with a reduction in homicide victimization rates, which were robust in multiple sensitivity analyses, more pronounced for firearm-related homicide deaths, and reduced homicide victimization inequities for Black Americans.

## Introduction

In the United States (US), homicide is a top 10 cause of death among people aged 5 to 54^1^ and therefore is a major contributor to early mortality and loss of societal potential. The upstream causes of homicide victimization, including concentrated poverty^2–4^, income inequality^5^, residential segregation^6,7^, and housing instability^8^, disproportionately harm racially minoritized people^9,10^. Consequently, homicide victimization is a major source of racialized inequities in life expectancy. Of all causes of death in the US, homicide victimization contributes the most person-years of life lost among Black people and more person-years of life lost among all racially minoritized people compared to White people^2,11–15^. Preventing homicide victimization is an urgent public health priority both in terms of reducing total life loss and reducing racialized inequities in life expectancy.

Despite the upstream causes of homicide victimization, approaches to reducing homicide have focused on individual- and community-level interventions. Many of these interventions are promising, including emergency department revictimization prevention interventions^16^, community-driven gang violence intervention programs^17,18^, and interagency collaborative approaches^19^. However, their effects are likely to be limited to the largely urban communities and individuals that directly receive interventions. To achieve sustainable reductions in homicide victimization, interventions that address the upstream societal and systemic determinants of violence are needed.

One upstream, state-level approach is increasing the minimum wage (MW) above the federally mandated $7.25 per hour. Depending on the increase in MW, this intervention can directly increase household income for more than 80 million hourly-waged workers in the US. While these policies have minimal antipoverty impacts when implemented alone^20– 22^, they appear to narrow racialized and gendered income inequalities^23,24^ and target the young adult age groups who disproportionately earn MW^25^ and are at greatest risk of homicide victimization^26^. As of 2024, 30 states had increased MW over the federal minimum of $7.25 per hour, prompting research on the health and quality of life impacts of MW increases.

MW increases are associated with many population health improvements, including fewer adolescent births^27^, lower infant mortality, especially among Black infants^28^, and lower premature mortality and increased life expectancy^29^. These health-protective effects have prompted calls from the American Public Health Association to increase the federal MW as a public health intervention^30^. Increasing MW has also been shown to reduce violence of various types, including violent externalizing behaviors in children^31^, child maltreatment particularly in very young children^32,33^, and alongside other household income support programs, reduced non-physical intimate partner violence^34,35^. However, existing studies typically do not examine MW increases’ effects on homicide specifically, nor do they evaluate the heterogenous effects of the policy on minoritized populations. One recent study evaluated the association between state-level Kaitz Index, or the ratio of the state median income to the MW as a measure of MW livability, and firearm homicide, finding that increased MW livability was associated with fewer firearm homicides^36^. However, this study did not compare the effects of MW increase itself in states with and without these changes.

We conducted a robust quasi-experimental study to evaluate the impact of state-level MW increases of at least $1.00 above the federal minimum on homicide mortality in the US between January 1, 2010, and December 31, 2019. We evaluate these effects for total homicide victimization mortality and disaggregated by firearm-involvement. We also examined heterogenous effects of MW increase by race, sex, and age strata to evaluate differential effects of the policy on subpopulations.

## Methods

We conducted controlled interrupted time series analysis (CITS) and synthetic controlled interrupted time series analysis (SCITS) to evaluate changes in homicide mortality trends within states that implemented MW increases to at least $8.25 per hour ($1 USD above the federal MW) between 2010 and 2019. We compared homicide mortality rates in states with MW increases to states that either never or always maintained a state MW of at least $8.25 during the study period. This study was reviewed and considered exempt by the Institutional Review Board at the University of North Carolina at Chapel Hill.

### Setting and Participants

US death records were obtained from the National Vital Statistics System (NVSS) to identify homicide deaths, firearm-involved homicides, and non-firearm-involved homicides (see outcomes). NVSS data also included age, sex, race, state, and county of decedents. The annual population of each state was derived from the NVSS bridged race postcensal state population estimates. The study period was from January 1, 2010, to December 31, 2019.

Information about state MW policy was acquired from the U.S. Department of Labor^37^. States that increased MW by at least $1 above the federal minimum of $7.25 between 2010 and 2019 were considered intervention states. There were 21intervention states including Arizona, Arkansas, California, Colorado, Delaware, Florida, Hawaii, Maryland, Massachusetts, Michigan, Minnesota, Missouri, Montana, Nebraska, New Jersey, New York, Ohio, Rhode Island, South Dakota, Vermont, and West Virginia (Figures S1 and S2).

The first pool of control states (“never-exposed”) consisted of those that never had a state MW of at least $8.25 during the study period. For ease of interpretation, we selected for inclusion in our main analysis the 13 states whose pre-intervention homicide mortality trends were parallel to that of the intervention states (Figure S2, Table S2). The second pool of control states (“always-exposed”) were those that always had a MW of at least $8.25 during the study period. We included the five always-exposed control states with parallel pre-intervention trends. Illinois was always exposed but was very influential to results due to a sharp and sustained increase in homicide rates in Chicago ^38^ coincident with the intervention time that were not reflective of overall national trends. Thus, despite having parallel pre-intervention rates, we excluded Illinois from the main analysis but included it in the sensitivity analysis. Further, for small subpopulations with differing trends from the total population (including Native and Asian people and children), we adjusted the pool of control states as described in Table S2. However, all candidate control states were included for the development of synthetic never-exposed and always-exposed controls during sensitivity analysis.

### Outcomes

Homicide mortality data for the United States was obtained from death certificate data from the National Vital Statistics System (NVSS). We used International Classification of Disease codes, 10^th^ revision (ICD10).to identify deaths due to homicide (U01-U02, X85-Y09, Y87.1), including firearm-involved homicides (U01.4, X93-X95) and non-firearm-involved homicide (all homicide ICD-10 codes not specific to firearms). Monthly homicide mortality rates per 100,000 person-years in each state were calculated by dividing the number of homicide deaths in a state and month by the annual population estimate in that state and multiplying by 12. For Asian and Native people there were zero homicides for some months due to small population, hence, we evaluated quarterly rather than monthly rates to avoid time periods with zero outcomes. Using these rates, we created time series of monthly (120 time points) and quarterly (24 time points) homicide mortality rates in all 50 US States.

### Exposure

The exposure of interest for this study was an increase in MW of at least $1.00 above the federal minimum during the study period from January 2010 to December 2019. Implementation time varied, with the earliest implementation in Vermont in January 2012 and the latest implementation in Missouri in January 2019 (Table S1). To examine the overall effect of MW increases in all 21 states together, we combined homicide mortality time series from all the intervention states by restructuring the calendar time. The month in which the MW increase intervention went into effect was time zero (‘0’), post-intervention time was “1, 2, 3….”, and pre-intervention time was “….-3, −2, −1.”

To set time zero among the control states for comparison purposes, we used the median and mode start time of MW increase in the intervention states, which was January 2016. The next most common month of intervention was January 2015, which we evaluated in sensitivity analysis (Appendix 1).

### Covariates

In death certificate data, individual race, ethnicity, sex, and age were available. Race was available as White, Black, American Indian/Alaska Native, Chinese, Japanese, Hawaiian, Filipino, other Asian, other non-White, or unknown race. Ethnicity options available include non-Hispanic, Cuban, Mexican, Puerto Rican, Central/South American, other Hispanic, or unknown. For compatibility with bridged-race population census estimates in the denominator, these were recoded as non-Hispanic White (hereafter White), non-Hispanic Black (Black), non-Hispanic American Native and Alaska Native (Native), non-Hispanic Asian and Pacific Islander (Asian), and Hispanic of any race (Hispanic). Death certificate and census sex were available as female and male. Death certificate and census age were categorized as under 18 years, 18 to 25, 26 to 35, 36 to 45, 46 to 55, 56 to 65, 66 to 75, and over 75 years. Due to the small number of homicide victimizations in individuals over age 55 years, we collapsed them into an age category of over 55 years. These covariates were used for stratified CITS analyses to evaluate effect measure modification.

### Statistical Analysis

We conducted CITS analysis using autoregressive moving average (ARIMA) models to evaluate changes in overall, firearm, and non-firearm homicide rate trends after implementation of state MW increases. The statistical models followed:

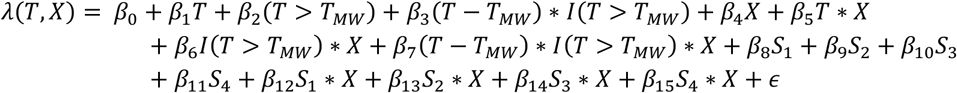

Where *λ*(*T, X*) is the homicide victimization mortality rate per 100,000 person-years at time interval *T* (months or quarters) for exposure group *X* (1=intervention, 0=control), *T*_*MM*_ is the month or quarter of MW increase for the intervention states and January 2016 for the control states, *I* is an indicator function that is 1 if the inside argument is true and otherwise 0, *S*_1−4_ are seasonality functions modeled with a harmonic time term with two sine/cosine pairs ^39^ to account for two annual homicide mortality peaks in winter and summer^40^, and *ϵ* is the error term. The auto-ARIMA function in the R forecasting package was used to identify the autoregressive, differencing, and moving average parameters for the models.

From the ARIMA models, the *β*_6_ represents the absolute change in homicide mortality rates in the intervention states in the first full month following the MW increase implementation compared to controls. *β*_7_ represents the sustained change in the trend of monthly or quarterly homicide mortality rates over the post-MW increase period in the intervention states compared to the trend change in controls. We calculated 95% confidence intervals (CIs) as a measure of precision, but these intervals should not be used to evaluate the existence of an effect^41^ since this study represents the complete source population of the United States. We then examined effect measure modification (EMM) by race and ethnicity, age, and sex groups to determine whether MW increases had differential subgroup effects.

### Sensitivity analyses

Given that there were few always-exposed states and those states were quite different from intervention states in terms of size, demographic composition, and political leaning (Table 1), we were concerned that the always-exposed control group may not be a good comparator and lacked exchangeability. Therefore, we developed synthetic controls representing the always-exposed and never-exposed states’ homicide mortality time series using the Synth package in R^42^ and then utilized them in SCITS analyses^43,44^. To build the synthetic controls, we utilized covariates representing state-level social determinants and policy environments to optimize comparability of pre-intervention trends among the intervention and control states. The covariates included time-varying state-level data encompassing the economic, policy, educational healthcare, neighborhood, and community social determinants of homicide mortality. These data and their sources are described in Table S3. Monthly covariates values were used where available, but most values were annual or semi-annual.

**Table 1.** Characteristics of study population (all 50 states) and analytic sample, by exposure group, pre- and post-intervention time.

| Variable | Pre-Intervention mean value |  |  |  | Post-intervention mean value |  |  |  |
| --- | --- | --- | --- | --- | --- | --- | --- | --- |
|  | Intervention group | Never-exposed control group | Always-exposed control group | All 50 states | Intervention group | Never-exposed control group | Always-exposed control group | All 50 states |
| Homicide mortality rate | 4.97 | 5.14 | 2.96 | 5.09 | 4.65 | 5.98 | 3.14 | 5.54 |
| Firearm-involved | 3.65 | 3.85 | 1.93 | 3.78 | 3.43 | 4.78 | 2.17 | 4.31 |
| Non-firearm-involved | 1.32 | 1.29 | 1.02 | 1.31 | 1.22 | 1.20 | 0.98 | 1.23 |
| % female | 50.9 | 50.7 | 50.4 | 50.8 | 50.8 | 50.7 | 50.3 | 50.7 |
| % in age group |  |  |  |  |  |  |  |  |
| Under 18 | 22.71 | 24.52 | 22.51 | 23.41 | 22.08 | 23.73 | 21.55 | 22.70 |
| 18 to 25 | 11.18 | 11.48 | 10.88 | 11.27 | 10.86 | 11.06 | 10.46 | 10.89 |
| 26 to 35 | 13.34 | 13.52 | 13.29 | 13.38 | 13.99 | 13.82 | 14.13 | 13.85 |
| 36 to 45 | 12.82 | 13.04 | 12.98 | 12.89 | 12.51 | 12.66 | 12.50 | 12.55 |
| 46 to 55 | 14.04 | 13.80 | 14.64 | 13.95 | 13.24 | 12.81 | 13.22 | 13.02 |
| Over 55 | 25.91 | 23.64 | 25.71 | 25.10 | 27.31 | 25.91 | 28.14 | 26.99 |
| % in race/ethnicity group |  |  |  |  |  |  |  |  |
| Asian | 6.98 | 3.60** | 6.81* | 5.24 | 8.99 | 4.22** | 7.92* | 6.43 |
| Black | 11.51 | 15.26 | 5.78 | 13.09 | 10.72 | 15.62 | 6.12 | 12.93 |
| Hispanic | 18.91 | 17.21 | 11.12* | 16.41 | 21.87 | 18.41 | 12.30* | 16.27 |
| Native | 0.73 | 0.72*** | 1.95* | 0.86 | 0.71 | 0.72*** | 1.92* | 0.85 |
| White | 61.88 | 63.20 | 74.35 | 64.4 | 57.72 | 61.04 | 71.73 | 61.52 |
| Covariates |  |  |  |  |  |  |  |  |
| % days poor air quality | 17.33 | 41.75 | 1.33 | 34.87 | 23.72 | 24.40 | 0.46 | 24.36 |
| % days high heat | 27.41 | 28.07 | 26.44 | 28.3 | 26.96 | 27.66 | 27.29 | 27.2 |
| % household without internet | 15.71 | 15.93 | 12.52 | 16.15 | 12.22 | 12.56 | 10.24 | 12.79 |
| % vacant housing units | 12.10 | 11.66 | 12.74 | 12.19 | 11.81 | 11.67 | 13.32 | 12.21 |
| % children within 1 mile of park | 57.83 | 50.52 | 58.89 | 52.31 | 74.22 | No data | No data | 74.22 |
| % children within 1 mile of school | 57.18 | 50.64 | 52.77 | 51.72 | 62.42 | No data | No data | 62.42 |
| Weighted mean SVI | 0.46 | 0.44 | 0.43 | 0.47 | 0.44 | 0.42 | 0.42 | 0.46 |
| Mean difference SVI | 0.75 | 0.86 | 0.70 | 0.78 | 0.76 | 0.85 | 0.68 | 0.79 |
| Hospital beds per 100k | 2.76 | 2.53 | 2.09 | 2.70 | 2.57 | 2.44 | 2.04 | 2.57 |
| Median income | 54,481.40 | 51,627.52 | 58,802.54 | 52,546.10 | 63,257.91 | 59,766.81 | 68,241.08 | 60,782.29 |
| Unemployment rate | 7.58 | 7.57 | 8.55 | 7.87 | 5.04 | 4.61 | 5.35 | 5.00 |
| Poverty rate | 14.30 | 15.09 | 12.90 | 14.85 | 12.47 | 13.05 | 11.05 | 12.94 |
| % HS graduate | 88.47 | 87.63 | 90.56 | 88.05 | 89.72 | 89.28 | 91.68 | 89.53 |
| Disability rate | 12.71 | 12.39 | 12.93 | 12.94 | 12.85 | 12.87 | 13.38 | 13.29 |
| Overdose rate | 13.72 | 13.26 | 13.46 | 13.70 | 19.92 | 20.49 | 20.78 | 19.74 |
| % schools with counselors | 79.71 | 85.54 | 73.84 | 82.35 | 82.52 | 85.85 | 71.38 | 83.40 |
| OOS days per 1k | 346.99 | 214.54 | 187.67 | 270.80 | 368.67 | 464.71 | 409.70 | 430.36 |
| School arrests per 1k | 1.12 | 1.32 | 0.91 | 1.28 | 1.73 | 2.47 | 1.98 | 2.12 |
| Total gun laws | 34.56 | 15.53 | 29.89 | 24.79 | 41.49 | 15.64 | 37.13 | 28.05 |
| Median (IQR) total gun laws | 24 (51) | 11 (10) | 22 (30) | 14 (19) | 30 (54) | 13 (9) | 36.5 (32) | 18 (30) |
| % with Medicaid expansion | 85.94 | 76.92 | 100 | 80.65 | 94.97 | 76.92 | 100 | 77.92 |
| % low income with low food access | 8.31 | 9.69 | 7.24 | 8.97 | 7.99 | 10.77 | 7.93 | 9.43 |
| % with low food access | 32.44 | 33.33 | 30.71 | 32.10 | 30.21 | 37.87 | 35.22 | 33.84 |

\*Oregon was included as an always-exposed control state for these populations, but not the main analysis. When excluding Oregon, the pre-intervention composition was 6.34% Asian, 4.94% Black, 11.38% Hispanic, 1.81% Native, and 75.53% White. The post-intervention composition was 7.35% Asian, 5.24% Black, 12.50% Hispanic, 1.79% Native, and 73.12% White.
\*\* In the pool of never-exposed control states for Asian people, the pre-intervention composition was 3.84% Asian and post-intervention composition was 4.48% Asian.
\*\*\* In the pool of never-exposed control states for Native people, the pre- and post-intervention composition was 0.85% Native.

## Results

Pre-and post-intervention homicide mortality rates, demographic composition, and social determinants covariate values demonstrated that intervention states were quite representative of patterns across all 50 states. When compared to the control states, we observed that intervention states were much more similar to never-exposed controls than to always-exposed controls (Table 1). The five always-exposed states had higher incomes, lower poverty, better educational attainment, lower social vulnerability and inequality in social vulnerability, and other determinants of their comparably low homicide mortality rates (Table 1). As such, the never-exposed control states represent a better comparator for drawing inferences about the effect of MW increases.

There were some important demographic differences between intervention and control states as well. Intervention states, especially post-intervention, had a large Hispanic population (21.87%) compared to always-exposed control states (12.30%), a Black population (10.72%) in between the never-exposed (15.62%) and always-exposed (6.12%) control states, and a White population (57.72%) lower than the never-exposed (61.04%) and always-exposed (71.73%) control states (Table 1). Further, Hispanic populations grew while White populations declined in all three groups of states from pre-to post-intervention. Meanwhile, Black populations declined in intervention states but grew in control states from pre-to post-intervention. These demographic differences, especially considering the racialized nature of homicide victimization, are important to note when evaluating EMM by race and ethnicity.

Compared to the never-exposed states, states that increased MW to at least $8.25 hourly had an immediate relative reduction in homicide mortality of −0.33 (−0.72, 0.07) per 100,000 PY in first month of the increase. This relative reduction was driven by a large absolute increase in homicide mortality in never-exposed control states as the intervention states also experienced a small absolute increase (Figure 1a). However, over the follow-up period, intervention states experienced a sustained annual decline of −0.22 (−0.37, −0.07) per 100,000 PY compared to never-exposed controls (Table 2).

**Table 2.**
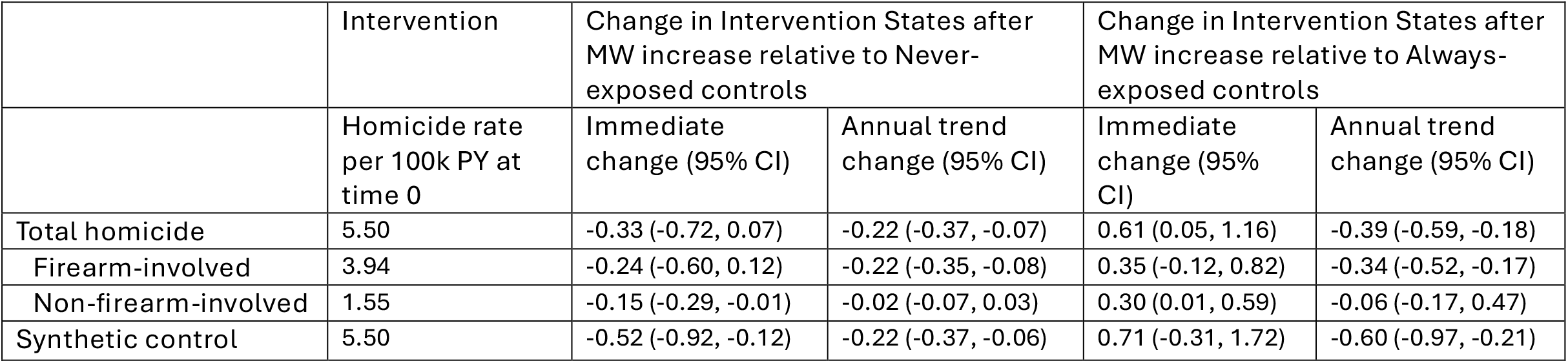
Immediate level change and sustained annual trend change coefficients and 95% confidence intervals for intervention states compared to never-exposed and always-exposed control states.

**Figure 1.**
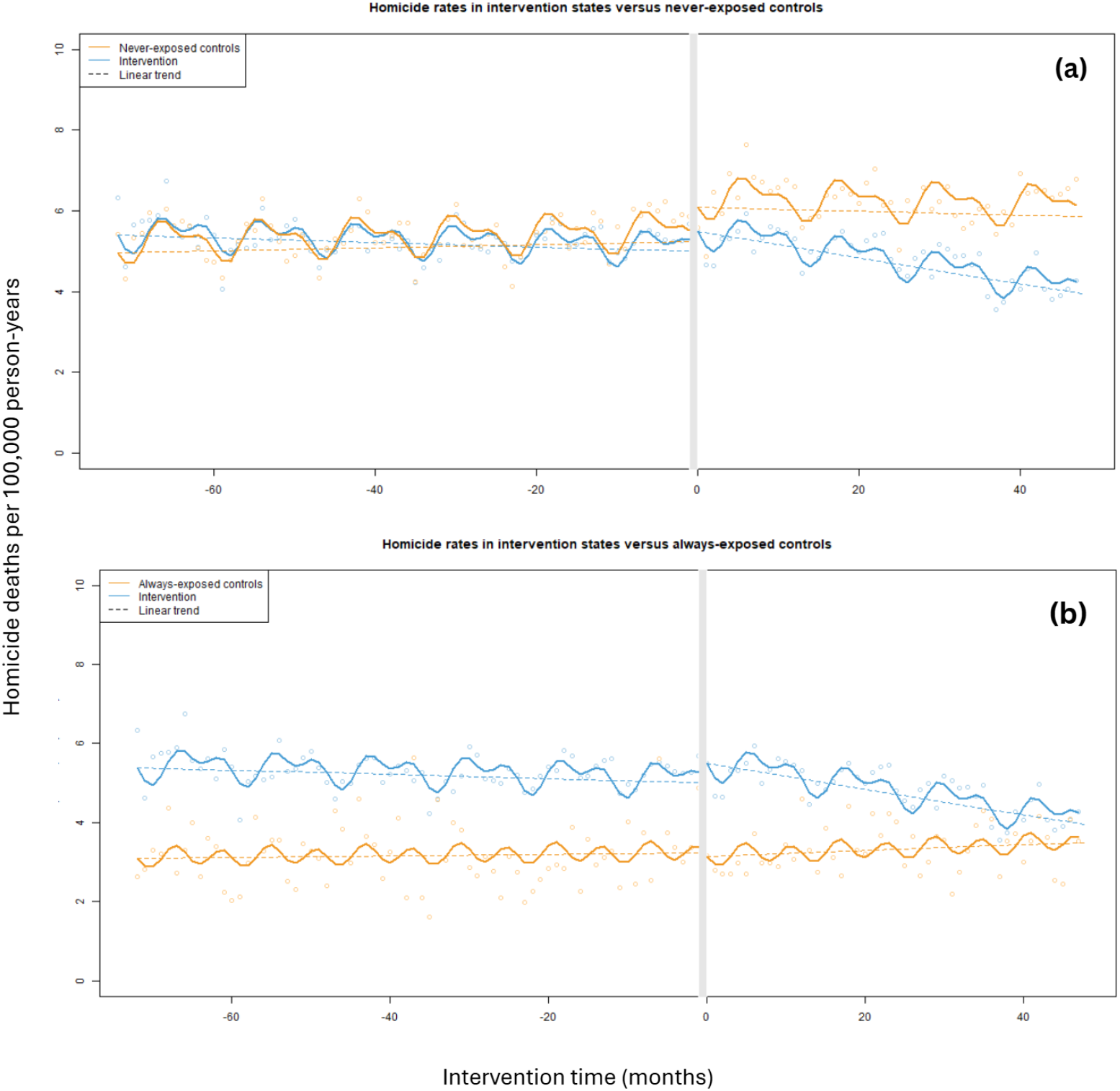
Total homicide rates in intervention states compared to states (a) never exposed to MW increase and (b) always exposed to MW increase.

Compared to always-exposed controls, MW increase states had an immediate relative increase in homicide mortality of 0.61 (0.05, 1.16) per 100,000 PY (Table 2), as the always-exposed control states did not experience notable immediate changes (Figure 1b). ^45^ However, there was then a sustained annual decline in MW implementation states of −0.39 (−0.59, −0.18) per 100,000 PY over the follow-up period (Table 2). Overall, homicide mortality rates in MW increase states were very similar to never-exposed controls in the pre-intervention period and then became more like always-exposed control states in the post-intervention period (Table 2, Figure 1).

Results by firearm involvement were also similar. As firearm-involved homicide mortality rates were higher than non-firearm-involved, the magnitude of immediate and sustained declines in firearm-involved homicide was greater than non-firearm-involved. The directions of effect were the same in both outcome types (Table 2).

Sensitivity analysis utilizing synthetic control states produced similar estimates, suggesting robustness of the main CITS results (Table 2; Appendix 1). Changing the Time ‘0’ for control states from January 2016 to January 2015 also produced similar results, especially with respect to never-exposed controls, suggesting robustness of the main analysis (Table S4; Figure S3).

In total, there were 30,852 homicide deaths (21,493 involving firearms) in intervention states during the post-intervention period. Using the level and slope change in the control states, we predicted counterfactual rates over the four years post-intervention to calculate the total number of lives saved after MW increase (Figure S4). A total of 5,657 fewer homicide deaths occurred in the intervention states after MW increases compared to the counterfactual given by the never-exposed states, and a total of 1,901 fewer homicide deaths after MW increase compared to the counterfactual given by the always-exposed states.

### Effect measure modification (EMM)

We found effect measure modification of the MW and homicide mortality association by race and ethnicity. While baseline homicide mortality rates suggest that Black and Native people disproportionately victimized (Table 3, Figures S5-S14) and the effect of MW increase on homicide mortality varied by race and ethnicity groups. Compared to never-exposed control states, all groups experienced an immediate decrease in homicide mortality rate. The magnitude of this decrease relative to baseline rates varied by group, with White and Asian people experiencing the largest declines (−10.7% and −20% respectively). Native people experienced the smallest decline at −1%, or −0.09 (−8.91, 5.76) per 100,000 PY. However, Native people experienced the largest sustained annual decreases in homicide mortality rate of any group at −17.2% or −1.58 (−8.91, 5.76) per 100,000 PY (Table 3). Compared to always-exposed states, the sustained annual decrease for Native people was −39.9%. We noted that these decreases were driven by avoidance of substantial homicide rate increases observed in control states rather than absolute rate declines in intervention states (Figures S8, S13).

**Table 3.** Immediate level change and sustained annual trend change coefficients and 95% confidence intervals for intervention states compared to never-exposed and always-exposed control states, by race/ethnicity, age, and sex strata.

|  | Intervention | Change in Intervention States after MW increase<br>relative to Never-exposed controls |  | Change in Intervention States after MW increase<br>relative to Always-exposed controls |  |
| --- | --- | --- | --- | --- | --- |
|  |  | Immediate change<br>(95% CI) | Annual trend change<br>(95% CI) | Immediate change<br>(95% CI) | Annual trend change<br>(95% CI) |
| Total | 5.50 | -0.32 (-0.72, 0.07) | -0.22 (-0.37, -0.07) | 0.61 (0.05, 1.16) | -0.39 (-0.59, -0.18) |
| Asian | 1.15 | -0.23 (-1.20, 0.74) | -0.07 (-1.15, 1.02) | -1.28 (-2.59, 0.04) | -0.81 (-2.29, 0.68) |
| Black | 23.74 | -1.32 (-3.10, 0.45) | -1.25 (-1.91, -0.58) | 1.40 (-2.34, 5.13) | -2.10 (-3.50, -0.70) |
| Hispanic | 5.00 | -0.28 (-1.01, 0.44) | 0.05 (-0.23, 0.32) | 0.001 (-1.31, 1.31) | 0.52 (0.03, 1.01) |
| Native | 9.18 | -0.09 (-6.44, 6.25) | -1.58 (-8.91, 5.76) | 2.60 (-4.41, 9.62) | -3.66 (-11.74, 4.42) |
| White | 2.62 | -0.28 (-0.56, 0.01) | 0.0001 (-0.11, 0.11) | 0.36 (-0.12, 0.83) | -0.13 (-0.30, 0.05) |
| Under 18 | 1.76 | -0.15 (-0.51, 0.21) | -0.14 (-0.27, -0.01) | 0.70 (-0.04, 1.45) | -0.04 (-0.32, 0.24) |
| 18 to 25 | 13.37 | -1.31 (-2.74, 0.13) | -0.72 (-1.26, -0.18) | 0.20 (-1.80, 2.19) | -0.68 (-1.43, 0.07) |
| 26 to 35 | 10.85 | -0.09 (-1.22, 1.05) | -0.22 (-0.64, 0.21) | 0.34 (-1.15, 1.82) | -0.84 (-1.40, -0.29) |
| 36 to 45 | 7.26 | -0.23 (-1.20, 0.74) | -0.41 (-0.77, -0.04) | 0.51 (-0.87, 1.89) | -0.40 (-0.91, 0.12) |
| 46 to 55 | 3.75 | -0.45 (-1.21, 0.32) | -0.01 (-0.30, 0.27) | 1.06 (-0.06, 2.17) | -0.62 (-1.04, -0.20) |
| Over 55 | 2.74 | -0.06 (-0.45, 0.34) | -0.14 (-0.29, 0.01) | 0.81 (0.13, 1.50) | -0.20 (-0.46, 0.05) |
| Male | 9.19 | -0.55 (-1.23, 0.13) | -0.36 (-0.62, -0.10) | 0.87 (-0.07, 1.81) | -0.63 (-0.98, -0.28) |
| Female | 1.92 | -0.11 (-0.38, 0.15) | -0.09 (-0.19, 0.01) | 0.40 (-0.01, 0.80) | -0.16 (-0.32, -0.01) |

Black people also experienced large sustained annual declines in homicide mortality rate of −5.3% or −1.25 (−1.91, −0.58) per 100k PY compared to never-exposed states and −8.8% or −2.10 (−3.50, −0.70) per 100k PY compared to always-exposed states. Unlike among Native people, these sustained decreases were driven by actual declines observed in intervention states (Figures S6, S11). In contrast, both White and Hispanic people experienced no sustained annual trend change in intervention states compared to never exposed control states, and Hispanic people experienced a large sustained annual increase in homicide mortality rate of 10.4% or 0.52 (0.03, 1.01) per 100,000 PY compared to always-exposed states (Table 3). This was driven by declines in homicide mortality among Hispanic people in always-exposed states that were not observed in intervention states (Figure S12). Asian people in always-exposed states experienced a large increase in homicide mortality at the intervention time followed by sustained increases which were not observed in intervention states (Table 3, Figure S10).

There was also some effect modification by age. While 18-to 25-year-olds had the highest homicide mortality rates at baseline, the immediate (−1.32 [−2.71, 0.07]) and sustained (−0.69 [−1.22, −0.17]) declines that they experienced compared to those in never-exposed states were larger than those experienced by other age groups (Table 3). Immediate increases and decreases compared to always-exposed states were variable, but every age group experienced sustained annual declines in homicide rate. There was no effect modification by sex, as immediate and sustained declines were proportional to baseline rates in each sex group.

## Discussion

Our study shows that state MW increases of at least $1.00 above federal MW were associated with sustained declines in homicide mortality compared to states that never increased MW, saving more than 5,000 lives from homicide death. Prior to MW increases, homicide mortality rates in intervention states were similar to those of states that never increased MW and were much higher than states that always had an increased MW. After MW increases, homicide mortality rates in intervention states declined, becoming more like states that always had an increased MW. Importantly, effect measure modification analyses showed that MW increase also reduced racialized inequities in homicide victimization for Black and Native people, through either substantial decrease of homicide mortality rates in intervention states or prevention of rate increases observed in control states. We also observed a larger rate decline in homicides among adults aged 18 to 25, who are disproportionately likely to be MW earners^25^. While the overall effect size is small, as would be expected with an upstream population health intervention^46^, that our findings were robust to sensitivity analysis, consistent across most subpopulations, and had a larger effect in those most exposed to violence and to earning MW suggests that MW increases are effective in reducing homicide.

These results align with the existing body of literature showing that MW increases produce both general population health improvements^28–30^ and reductions in certain measures of violence^31,32,34,35^. This is because MW increases reduce social determinants such as income inequality^22,43,44^ and racialized gaps in income and poverty,^47,48,49,50^, thereby improving household financial security ^51,52^ and reducing housing instability^53–55^. Our findings also support research showing that improvements in MW livability are associated with reductions in firearm homicide victimization particularly for racially minoritized people^5^. MW increases may also have a direct individual-level effect for individuals engaged in MW employment by improving individual predictors of violence victimization, like household financial security ^51,52^ and housing instability^53–55^.

Despite these population benefits, many individuals in poverty and at high risk of homicide victimization - including disabled people^56–58^, people experiencing intimate partner violence^59^, people with criminal legal involvement^60–62,^ and people experiencing houselessness^54,63,64^ – may not be able to participate in the labor workforce and thus MW policy does not directly change their circumstances. Lack of workforce participation could be one reason why we observed no effect modification by sex even though a greater share of females earn MW^25^, and both low income and poverty predict intimate partner homicide victimization among females^65^. In part, economic dependence may act as a barrier to leaving violent relationships. Unfortunately, those.most at risk of intimate partner homicide may be less likely to benefit from MW increases^66^ and other earned income supports like EITC^67,68^ due to lack of formal work^69^. Additional anti-poverty interventions not tied to workforce participation are necessary to reduce the risk of violent death among these vulnerable populations.

Our findings are supported by the strength of the controlled interrupted time series design, which utilized multiple pools of real and synthetic controls to account for secular trend and control potential confounders^44^. Further, our analysis included all available homicide mortality data via NVSS death certificate records from all 50 U.S. states over a 10-year period, permitting estimation of effects in under-researched populations like Asian and Native people.

We recognize that there are some limitations brought by the data. First, cause of death and race and ethnicity misclassification on death certificates are barriers to accurate homicide mortality rate estimation. Second, the always-exposed control group was small and non-comparable due to the reality of state-level MW policy making in the US. We addressed this limitation by using a synthetic always-exposed control state, but a future replication study that shifts some early adopting intervention states into the always-exposed group and follows up beyond the year 2020 will be an important addition to this work. Third, minimum wage increases are not randomly assigned, these policy actions may be correlated with other policies affecting homicide rates. However, the overlapping similarity between the intervention and never-exposed states and the use of synthetic controls address this concern. Finally, we did not evaluate effects past 2020 due to the complex intersection of COVID-19 homicide mortality effects, economic impact payments, and MW policy increases which merit a separate evaluation and will be addressed in future research.

## Conclusions

This quasi-experimental study showed that states that increased MW to at least $1 USD above the federal minimum experienced sustained declines in homicide mortality rates, especially compared to states that were never exposed to an increased MW. MW increases and reduction of racialized inequities in homicide victimization for Black and Native people. Future studies should evaluate whether and how these effects were sustained or changed by the COVID-19 pandemic and its economic interventions.

## Data Availability

All data produced in the present work are contained in the manuscript.

