## Supplementary material for "Impact of minimum wage increases on homicide mortality in the US": CITS-MinWage-Homicide-Supplement-Update_2026-05-21.docx

**Table S1. Month and year of state MW increase for intervention states.**

| State | Implementation date |
| --- | --- |
| AZ | 01/2017 |
| AK | 01/2017 |
| CA | 07/2014 |
| CO | 01/2016 |
| DE | 01/2015 |
| FL | 01/2018 |
| HI | 01/2016 |
| MD | 01/2017 |
| MA | 01/2015 |
| MI | 01/2016 |
| MN | 08/2015 |
| MO | 01/2019 |
| MT | 01/2018 |
| NE | 01/2016 |
| NJ | 01/2014 |
| NY | 01/2015 |
| OH | 01/2018 |
| RI | 01/2015 |
| SD | 01/2015 |
| VT | 01/2012 |
| WV | 01/2016 |

**Figure S1. Flowchart of selection of states for inclusion in analysis.**

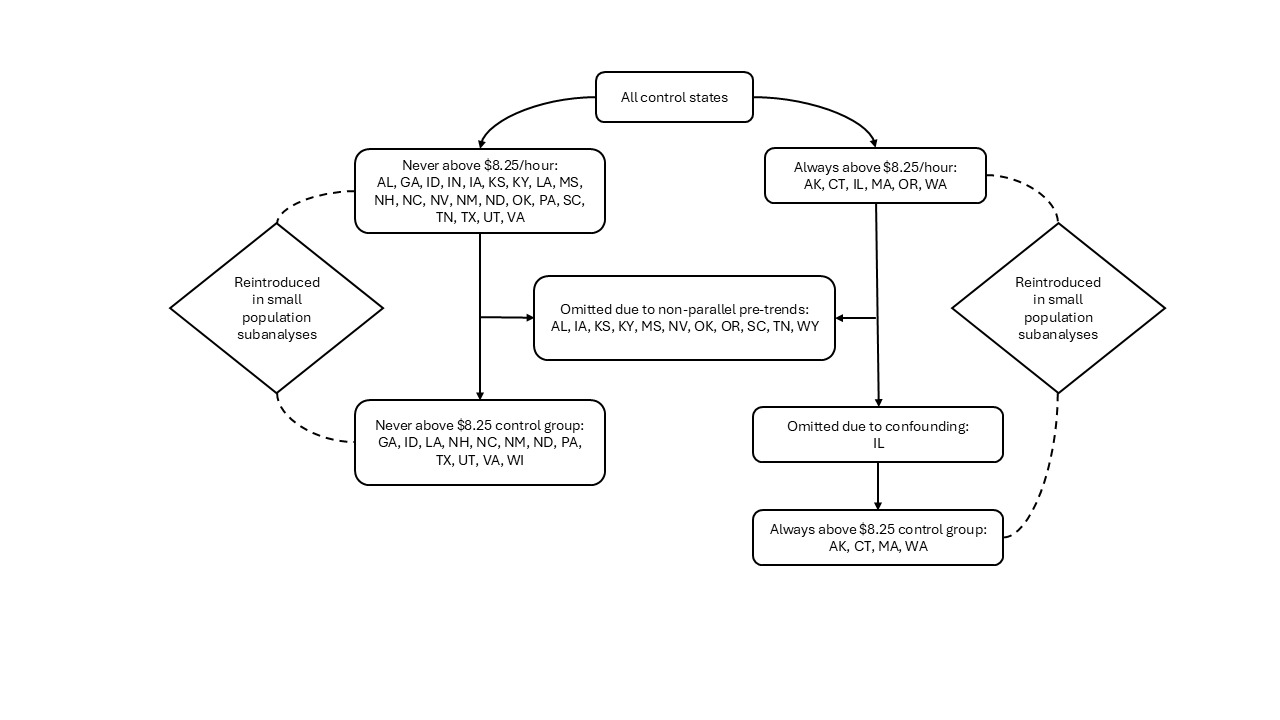

**Figure S2. Map of states included in analytic sample, by MW exposure group.**

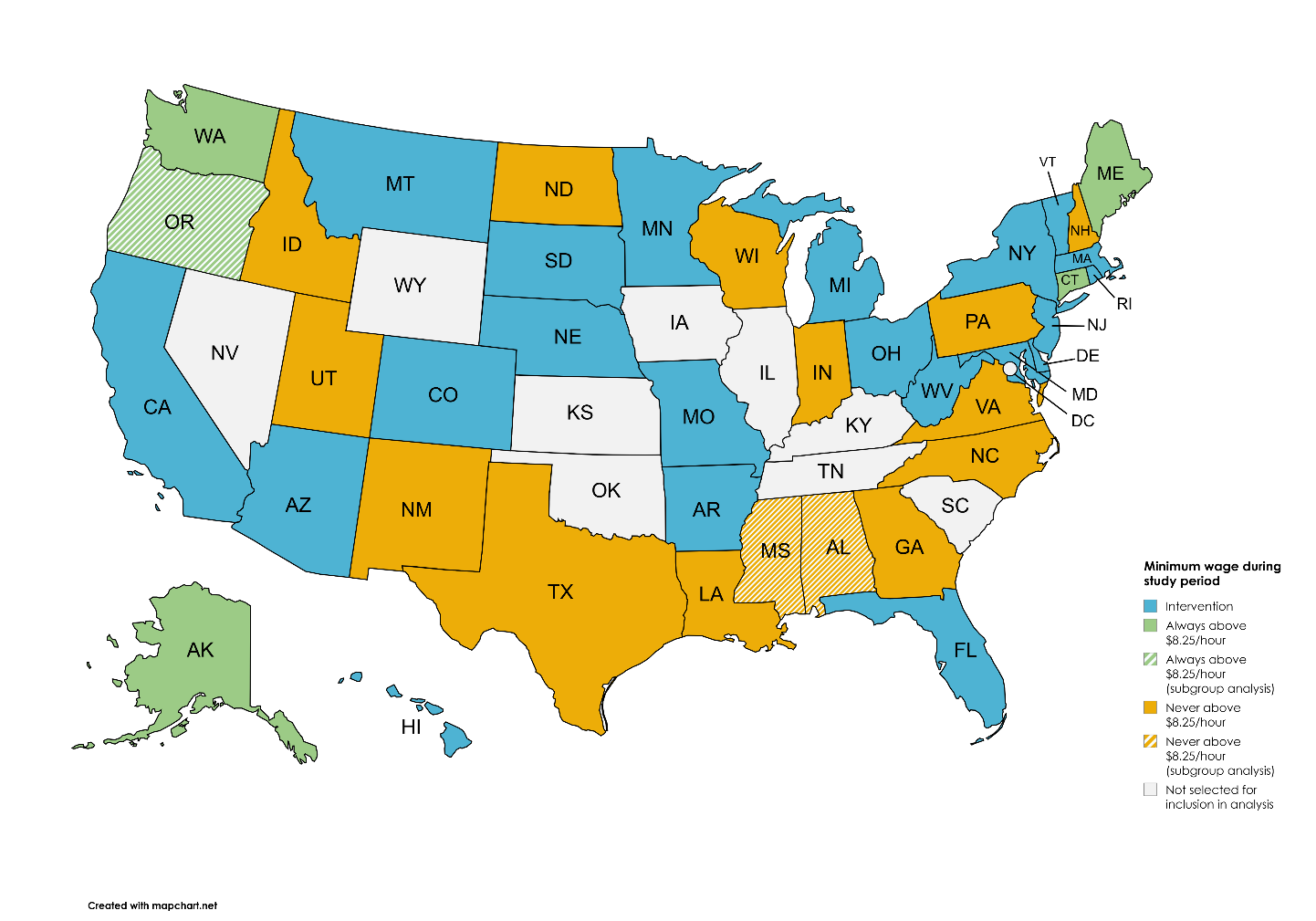

**Table S2. States included in analytic sample by outcome and subpopulation**

|  | Never-exposed states  (Never had minimum wage of at least $8.25 between 2010-2019) | Always-exposed states  (Always had minimum wage of at least $8.25 between 2010-2019) |
| --- | --- | --- |
| All candidate states | AL, GA, ID, IN, IA, KS, KY, LA, MS, NH, NC, NV, NM, ND, OK, PA, SC, TN, TX, UT, VA, WI, WY | AK, CT, IL, ME, OR, WA |
| Main analytic sample | GA, ID, IN, LA, NH, NM, NC, ND, PA, TX, UT, VA, WI | AK, CT, ME, WA |
| Asian | LA, MS, NC, PA, TX, UT, VA | AK, CT, ME, OR, WA |
| Black | Main analytic sample | Main analytic sample |
| Hispanic | Main analytic sample | AK, CT, ME, OR, WA |
| Native | AL, GA, ID, LA, MS, NH, NC, ND, PA, UT, VA, WY | AK, CT, ME, OR, WA |
| White | Main analytic sample | Main analytic sample |
| Under 18 | ID, IN, NH, NM, NC, ND, PA, TX, UT, VA, WI | Main analytic sample |
| 18 to 25 | Main analytic sample | Main analytic sample |
| 26 to 35 | Main analytic sample | Main analytic sample |
| 36 to 45 | Main analytic sample | Main analytic sample |
| 46 to 55 | Main analytic sample | Main analytic sample |
| Over 55 | Main analytic sample | Main analytic sample |
| Female | Main analytic sample | Main analytic sample |
| Male | Main analytic sample | Main analytic sample |

**Appendix 1. Sensitivity Analyses**

**1.1 Synthetic controls**

Synthetic control never-exposed and always-exposed states were constructed with the R Synth package^[1]^ using NVSS homicide mortality rates and social determinants of health covariates (Table S3). This package weights covariates and states to construct a maximally comparable synthetic state that we then used in CITS modeling. The never-exposed synthetic control state consisted of 27.4% TX, 25.7% PA, 12.4% NC, 9.5% VA, 6.4% AL, and 5.2% UT. All other states contributed less than 5% to the synthetic state. The always-exposed synthetic control state consisted of 52.6% CT, 30.9% AK, 15.4% WA, 1.1% OR, and 0% ME. Compared to our real state control models, the immediate change compared to synthetic never-exposed control states was larger in magnitude, but in the same direction of effect. The annual trend change in synthetic never-exposed controls was exactly the same as in real never-exposed controls, suggesting that our real state model was an excellent counterfactual for sustained trends. Immediate and annual trend changes in synthetic always-exposed controls were larger in magnitude but in the same direction as our real state analysis. Given these results, we concluded that our main analysis was robust.

**Table S3. Social determinants of health covariates, data years, and sources for construction of synthetic control states.**

| Median household income | ACS (2000 – 2019) |
| --- | --- |
| Unemployment rate | ACS (2000 – 2019) |
| Percent under poverty line | ACS (2000 – 2019) |
| Percent high school graduate | ACS (2000 – 2019) |
| Disability rate | ACS (2000 – 2019) |
| Percent population institutionalized | ACS (2000 – 2019) |
| Percent with SNAP/cash assistance | ACS (2000 – 2019) |
| Percent of schools with school counselors | CRDC (2011 – 2017) |
| Days lost to out-of-school suspension per 1,000 enrolled students | CRDC (2011 – 2017) |
| In-school arrests per 1,000 enrolled students | CRDC (2011 – 2017) |
| Medicaid expansion | Medicaid.gov (2013 – 2020) |
| Child enrollment in Medicaid and CHIP | Medicaid.gov (2013 – 2020) |
| Per-capita hospital beds | Kaiser Family Foundation (2000 – 2019) |
| Age-adjusted opioid overdose mortality rate | CDC WONDER (2000 – 2019) |
| Percent children with 1-mile access to public park | CDC NEPHTN (2010, 2015) |
| Percent children with 1-mile access to school | CDC NEPHTN (2020, 2015) |
| Percent households with home internet access | CDC NEPHTN (2001 – 2018) |
| Percent days with poor air quality | CDC NEPHTN (2001 – 2016) |
| Percent days > 90^th^ percentile heat | CDC NEPHTN (2013 – 2018) |
| Percent vacant housing units | CDC NEPHTN (2009 – 2018) |
| Mean social vulnerability index | CDC NEPHTN (2010 – 2018) |
| Max – min social vulnerability index | CDC NEPTHN (2010 – 2018) |
| Percent low income and low food access | USDA (2015, 2019) |
| Percent low food access | USDA (2015, 2019) |

**1.2 Intervention time**

**Table S4. Immediate and sustained trend changes in homicide mortality after minimum wage increase, main analysis and sensitivity analyses**

|  | **Immediate change (95% CI)** | **Annual trend change (95% CI)** |
| --- | --- | --- |
| *Never exposed, main analysis* | -0.32 (-0.72, 0.07) | -0.22 (-0.37, -0.07) |
| *Always exposed, main analysis* | 0.61 (0.05, 1.16) | -0.39 (-0.59, -0.18) |
| Never exposed, 2015 interruption | -0.38 (-0.83, 0.06) | -0.24 (-0.42, -0.06) |
| Always exposed, 2015 interruption | -0.07 (-0.63, 0.49) | -0.23 (-0.45, -0.01) |
| Always exposed + Illinois, 2016 interruption | -0.86 (-1.44, -0.29) | -0.08 (-0.30, 0.13) |
| Always exposed + Illinois, 2015 interruption | -0.51 (-1.17, 0.16) | -0.34 (-0.60, -0.08) |

Sustained trend change results when time zero was set to January 2015 were similar to main results. Immediate level changes were similar to main analysis when evaluating intervention states compared to never-exposed controls, but changed direction when compared to always-exposed controls (Table S4). This is likely due to greater variability in homicide mortality rates in the smaller group of always-exposed control states. We concluded that our findings that minimum wage increases were associated with sustained declines in homicide mortality rates compared to control states were robust to intervention time, but cautious interpretation of level changes compared to always-exposed states is warranted.

**Inclusion of Illinois**

Results when including Illinois in the always-exposed control states with a 2016 interruption time were quite different than when excluding it, due to a notable spike in homicide rates in Chicago, IL coincident with time zero. This spike in homicide mortality was driven by several factors including strained community relations and a budget impasse causing interruptions to violence prevention programs and poverty relief services^[2]^. Inclusion of this time- and place-specific confounding event resulted in a reversal of effect compared to what was observed when Illinois was excluded from the main analysis (Table S4, Figure S3). This effect reversal was still seen, but to a much less extreme extent, when the control state interruption time was set to January 2015. Given that this confounding event was not representative of homicide mortality patterns observed in the remaining always-exposed control states, we concluded that excluding Illinois provided a more generalizable result, though all results are presented here for transparency.
**Figure S3. Homicide mortality rates in intervention states versus always-exposed control states including Illinois.**

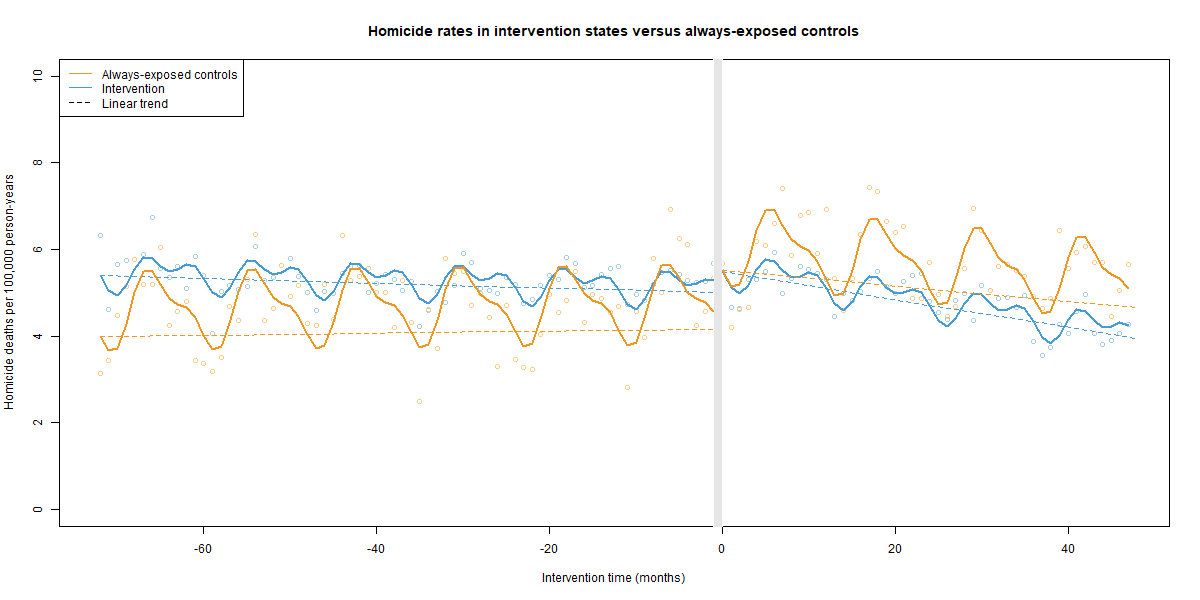

**Figure S4. Counterfactual rates for intervention states given by (a) never-exposed states and (b) always-exposed states’ post-intervention level and slope changes.**

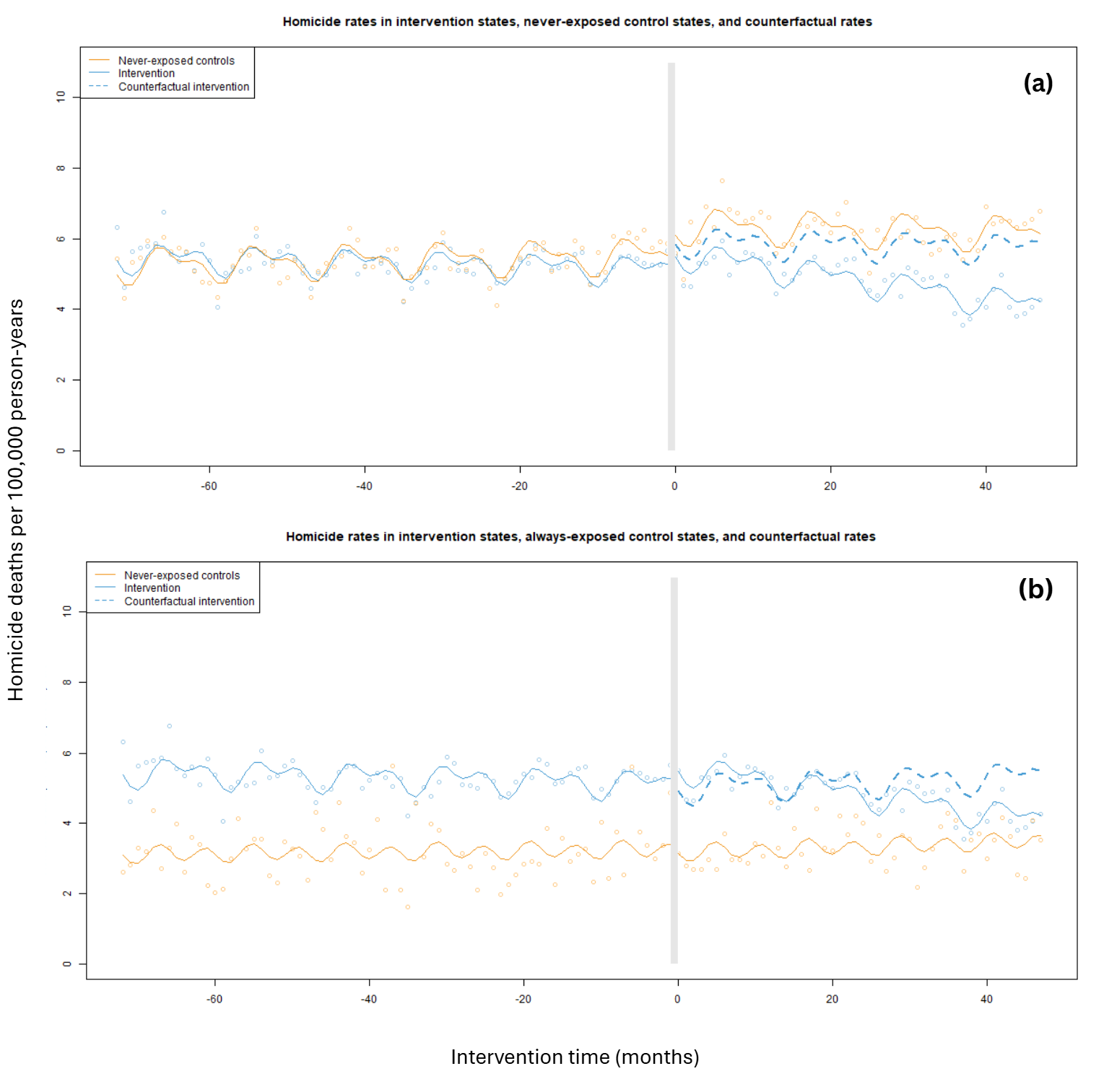

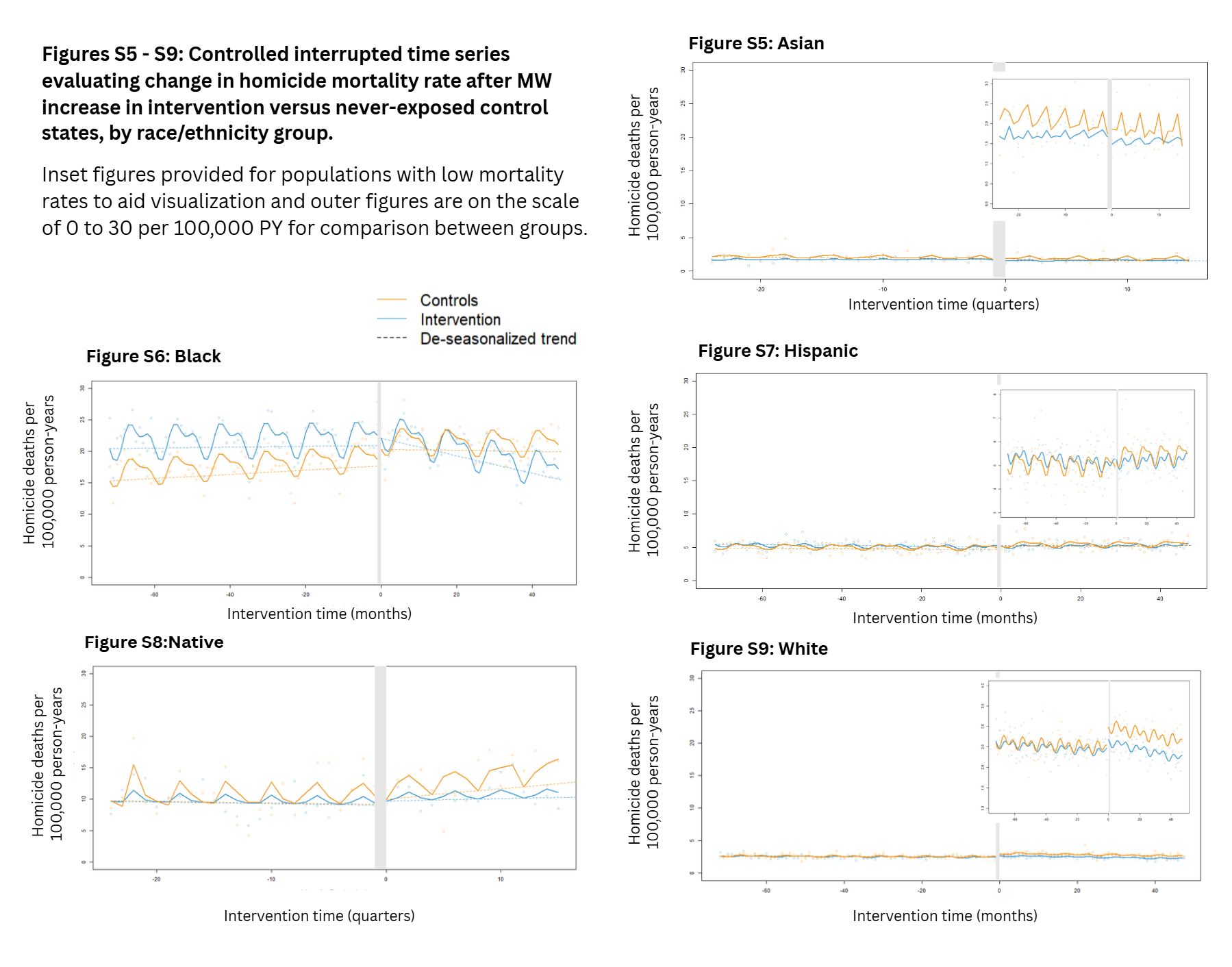

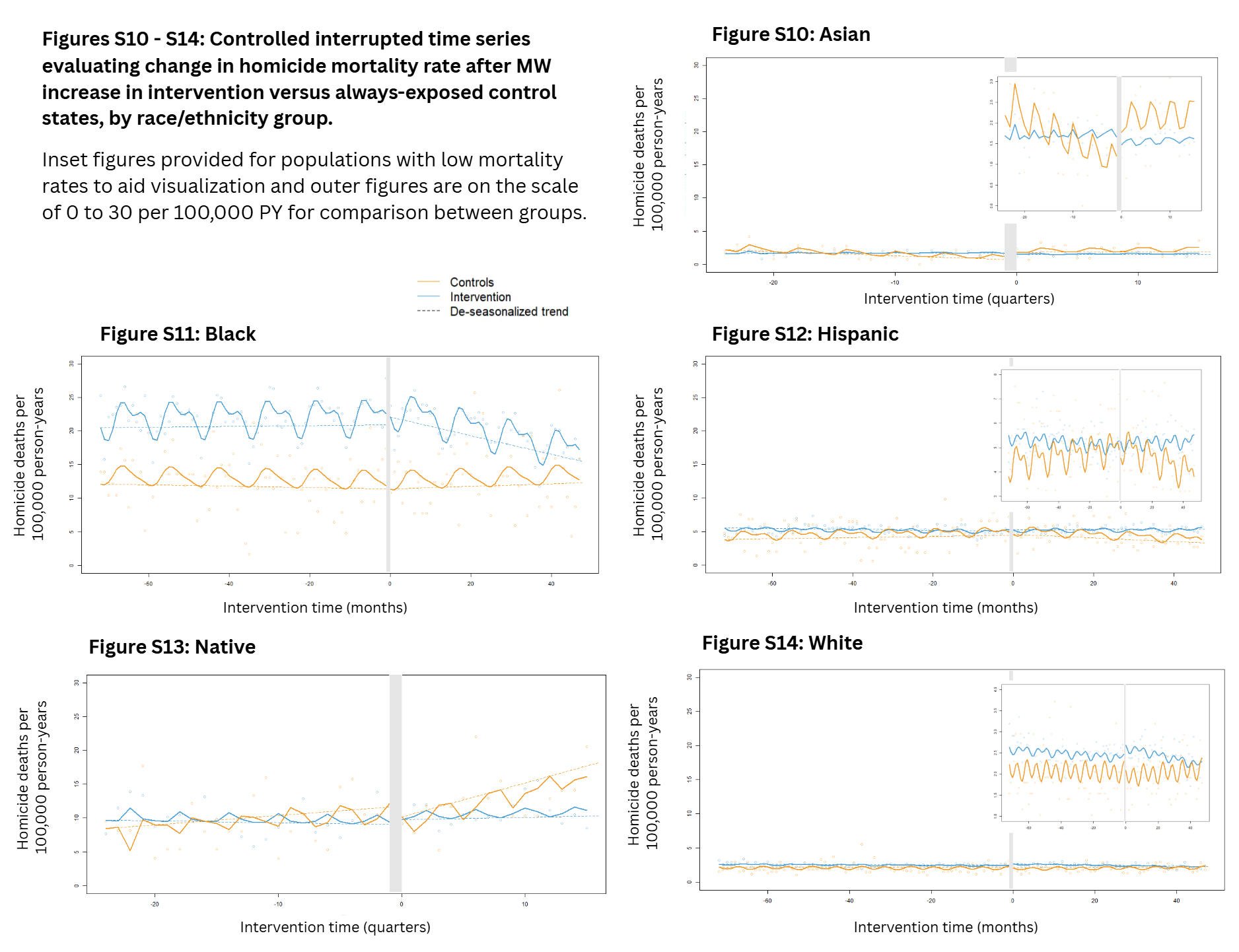

1. Abadie, A., Diamond, A., & Hainmueller, J. (2011). Synth: An R Package for Synthetic Control Methods in Comparative Case Studies. *Journal of Statistical Software*, *42*(13), 1–17. <https://doi.org/10.18637/jss.v042.i13>
2. Dastoor JD, Thomas A, Slocum JD, Regan S, Stone L, Richardson JB, Mason M, Johnson JK, Lin K, Stey A. Investigating the 2016 surge in firearm violence in Illinois, USA, through community-based organisations: a qualitative study. Inj Prev. 2024 Nov 21;30(6):503-508. doi: 10.1136/ip-2023-045075. PMID: 38448213; PMCID: PMC11377856.
